# A quantitative universal NGS-based ctDNA assay for hepatoblastoma

**DOI:** 10.1101/2022.09.20.22279947

**Authors:** Smadar Kahana-Edwin, James Torpy, Lucy E. Cain, Anna Mullins, Geoffrey McCowage, Sarah E. Woodfield, Sanjeev A. Vasudevan, Dan P.T. Shea, Andre E Minoche, Sarah Kummerfeld, Leonard D. Goldstein, Jonathan Karpelowsky

**Author notes:** Corresponding author: Smadar Kahana-Edwin, Kerry Packer Building, Cnr Hawkesbury Road and Hainsworth Street, The Children’s Hospital at Westmead, Westmead NSW 2145, Australia. These authors contributed equally to this work (Smadar Kahana-Edwin, James Torpy, Lucy E. Cain, Anna Mullins).

## Abstract

Driver mutations in *CTNNB1* are a hallmark of hepatoblastoma and offer a common biomarker for a liquid biopsy approach based on the presence of *CTNNB1* circulating tumor DNA (ctDNA). We developed and investigated the utility of a quantitative universal next-generation sequencing (NGS) ctDNA assay for hepatoblastoma (QUENCH) to detect *CTNNB1* ctDNA and assessed the links between ctDNA and current clinical indicators/biomarkers in hepatoblastoma. Applied to patients with hepatoblastoma, we demonstrate quantitation of various variants including single base substitutions and deletions down to 0.3% variant allele frequency, with 65% sensitivity and 100% specificity at the patient level, to allow biopsy-free tumor genotyping and sensitive ctDNA quantitation. CtDNA positivity correlates with tumor burden and ctDNA levels correlate with macroscopic residual disease and treatment response, thus providing promising evidence for the utility of quantitative ctDNA detection in hepatoblastoma.

## INTRODUCTION

Hepatoblastoma is the most common liver tumor diagnosed in children. It occurs predominantly in young children under the age of 3 years and has a rising incidence. Hepatoblastoma is generally managed with pre- and/or post-operative chemotherapy, tumor resection with partial hepatectomy, or in selected cases, liver transplantation ^1^. Risk assessment and subsequent therapy planning are essential to achieve good outcomes for this disease. To address this, international risk classification standards were created based on the PRE-Treatment EXTent of tumor (PRETEXT) imaging staging system ^2^, serum Alpha-fetoprotein (AFP) values, and patient’s age to separate patients into very low risk, low risk, moderate risk, and high risk ^3^ groups. Survival rates have significantly improved in children with hepatoblastoma, especially in the high risk group. Contributors to improved survival include the use of risk-adapted chemotherapy and advanced surgical approaches such as indocyanine green (ICG) fluorescence-guided surgery and liver transplantation ^4^. However, about 12% of hepatoblastoma patients who have achieved a complete remission are likely to relapse in the liver and/or lungs ^1^, and could benefit from better measurable residual disease (MRD) detection and modification of treatment options. Furthermore, some of the most high risk tumors, especially relapse tumors, do not secrete AFP and thus lack the AFP biomarker^5^. By contrast, this risk stratification system is lacking good efficacy to distinguish between the prognosis of the very low risk group and low risk group ^6^ and could benefit from further development. This is needed as distinction between low risk and very low risk groups determines the use of adjuvant chemotherapy post-operatively in the former or observation only for the latter, which is very important in the context of morbidity associated with platinum-based chemotherapy.

Cell-free circulating tumor DNA (ctDNA) is a surrogate for the tumor genome and is frequently the most accessible and least invasive clinical sample for applications such as therapy selection, post-treatment monitoring, and early cancer screening - also known as cancer liquid biopsy ^7,8^. Hepatoblastoma shows one of the lowest mutational burdens among all cancers (0.52/Mb on exonic regions), having recurrent mutations associated with exon 3 of the *CTNNB1* gene in 80-100% of sporadic tumors ^9–11^. We previously reported an association between *CTNNB1* ctDNA variant allele frequency (VAF) identified through digital droplet PCR (ddPCR) and levels of serum AFP over the course of treatment of three patients with hepatoblastoma ^12^. However, applying tumor-informed custom probe designed ddPCR assays poses a challenge for real-time data at diagnosis. This is because the location and length of mutation can vary from patient to patient, information that can only be determined by sequencing of tissue biopsies.

Due to the rarity of ctDNA and the dilution effect of non-malignant cell free DNA (cfDNA), high assay sensitivity and quantitative ctDNA assessment are required to provide an accurate status of disease burden. Here, we developed a **q**uantitative **u**niv**e**rsal next-generation sequencing (**N**GS) **c**tDNA assay for **h**epatoblastoma (QUENCH) to allow a tumor-agnostic near real-time evaluation of ctDNA. Our primary objective was to investigate the utility of QUENCH to detect *CTNNB1* ctDNA. Our secondary objective was to compare the levels of (1) QUENCH ctDNA, (2) ddPCR ctDNA, and (3) AFP, and verify if they correlate with tumor burden and treatment response. We applied QUENCH to a cohort of 38 samples from 20 patients and demonstrated that *CTNNB1* ctDNA variants were accurately quantified down to 0.3% VAF, including single-base substitutions and deletions, and that VAF levels correlated with tumor burden and dynamic treatment response.

## RESULTS

### Characteristics of the Study Population

Twenty patients were included in this study with the following features: patients were diagnosed with hepatoblastoma at a median age of 24 months (range 3 to 171), with a mean AFP level at diagnosis of 140,223 kIU/L (n=10). Eight of 20 (40%) patients were female, 6 of 20 (30%) had metastatic disease at diagnosis, and the majority (9 of 11 with available data, 82%) demonstrated mixed epithelial and mesenchymal histology.

Somatic mutations in the exon 3 region of the *CTNNB1* gene were evaluated with Sanger sequencing of the primary tumors or metastases as previously described ^12^. Data was available for 18 of the 20 patients to demonstrate 7 missense single nucleotide variants (SNVs), 10 deletions ranging from 10 to 348 bp, and 1 wild type *CTNNB1* (Supplementary Materials Fig S1).

**Figure S1.**
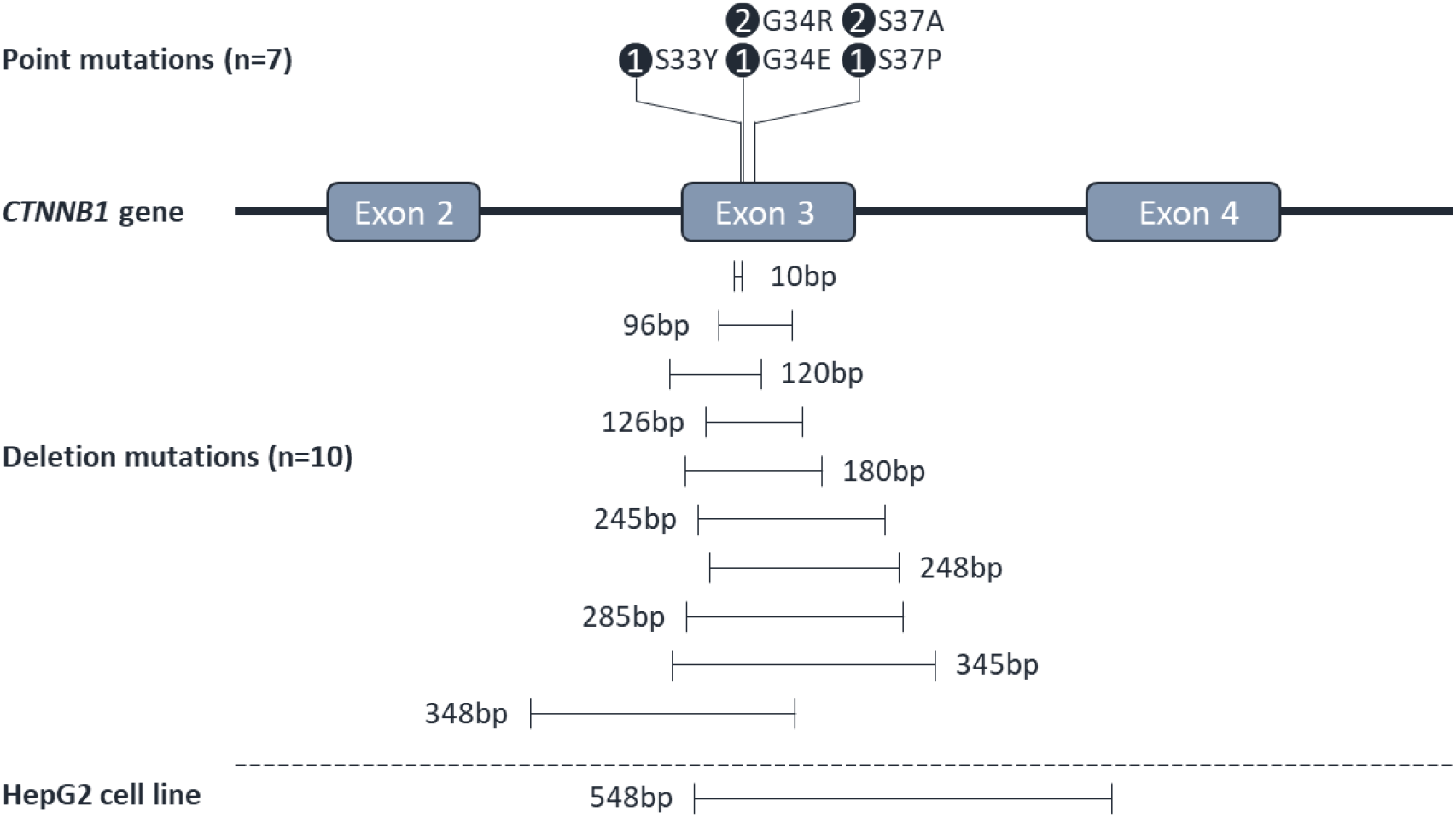
Schematic view of the *CTNNB1* exon-intron structure targeted by QUENCH and the mutations in the studied cohort and HepG2 cell line. Pathogenic SNV missense mutations (in circle - the number of patients with the same mutation) and SV deletion mutations identified in the study cohort are shown.

### Assay Sensitivity at Low VAF Levels

QUENCH development was based on UMI incorporation as well as single primer extension capture. In addition to error-correction, UMI allows more accurate determination of ctDNA levels from VAF. A customized in-house structural variant (SV) calling method and software was developed to detect deletions in *CTNNB1* from short DNA sequences such as cfDNA (see Methods).

The scarcity of ctDNA and the extremely low amount of cfDNA (in the ng range) might hinder successful sampling of rare variants. In order to develop and benchmark our variant caller to detect low fraction somatic variants in plasma, precision, accuracy, and linearity were determined at low variant levels. For precision and accuracy, we created a reference standard DNA with defined variants at low allele fractions (see Methods). We used 20 ng input DNA to reflect the typical analysis of 0.5 mL plasma from patients with hepatoblastoma (see chapter below), and sequenced to a median average read depth of 627x (407-837x) and median average UMI depth of 307x (221-430x). The LoD was determined as the lowest VAF that could be detected with a minimum 1 variant copy for SV calling. Given that 20 ng DNA represents about 3,030 diploid human genome equivalents ^13^, the maximum expected sensitivity was 0.03% VAF. Accordingly, we observed a linear detection of the variant down to VAF of 0.1% (R^2^ = 0.9932), however, were not able to detect the variant at 0.03% or beyond that level (Fig 1), setting the LoD at 0.1% VAF.

**Figure 1.**
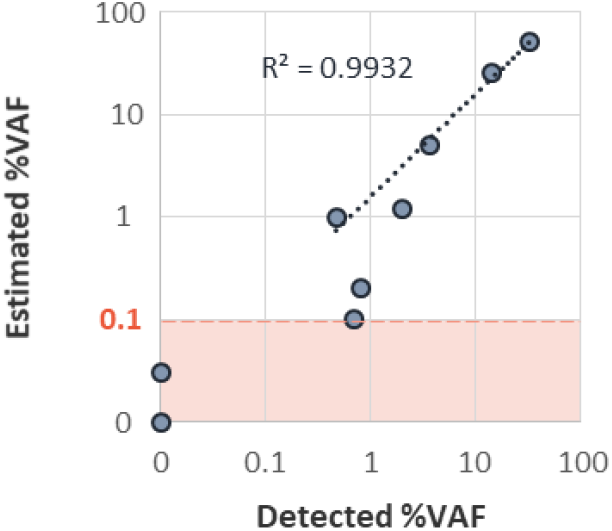
QUENCH limit of detection and linearity. Genomic DNA of the hepatoblastoma cell line HepG2 with known CTNNB1:c.73_420 deletion was diluted in normal *CTNNB1* genome to generate VAF titration series of 0%, 0.03%, 0.1%, 0.2%, 0.3%, 1%, 1.2%, 5%, 25%, and 50%. Theoretical (Y axis) and QUENCH-detected (X axis) VAF are plotted. Linear regression is presented for VAF ≥ 0.1%.

To provide a QUENCH false-positive call rate, the LoB was determined using four samples with non-mutated *CTNNB1*. No variants were detected in any of the four samples, thus no QUENCH false-positive call was made, making the LoB 0% VAF.

### Assay Verification - Concordance with ddPCR

To validate QUENCH quantitation capability and evaluate its sensitivity in clinical samples, we compared QUENCH VAF with VAF from ddPCR. Twenty-three samples were evaluated from 8 patients, 4 harboring SNVs and 4 with deletions in their primary tumors (Fig 2). QUENCH and ddPCR had excellent concordance in 10 samples with VAF above 0.35% (R^2^ = 0.9606, Fig 2a). However, QUENCH sensitivity was inconsistent in ddPCR VAF between 0.1% and 2.2%, as only 4 samples were detected by QUENCH while 6 additional samples were detected by ddPCR, even though the theoretical LoD demonstrated above was 0.1%. No sample was QUENCH-positive and ddPCR-negative, and 6 samples were negative by both QUENCH and ddPCR. Compared with ddPCR, QUENCH showed sensitivity of 59% and specificity of 100% (Fig 2b).

**Figure 2.**
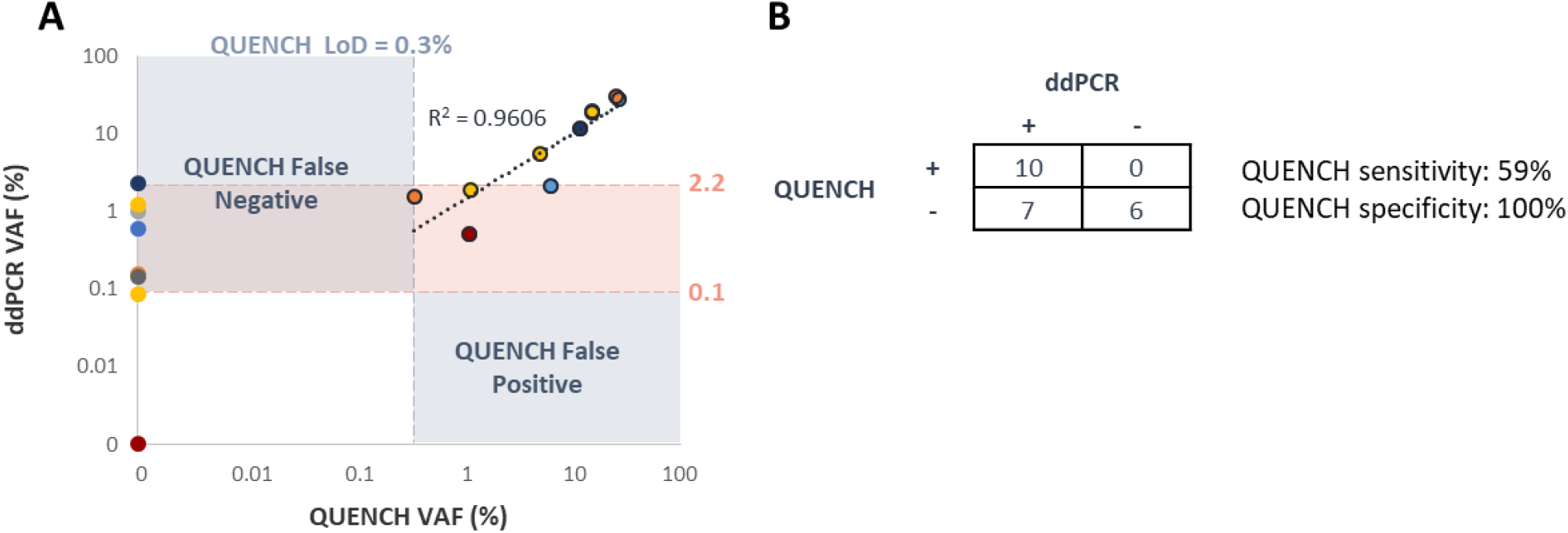
QUENCH verification - concordance with ddPCR. Matched ddPCR and NGS results from 8 patients (color-coded according to each patient). **(A)** High concordance (R^2^=0.9606) observed in samples with QUENCH VAF > 0.3%. ddPCR shows higher sensitivity for VAF < 2.2%. **(B)** Sensitivity, specificity, and concordance of variant detection evaluated by QUENCH and ddPCR.

### CtDNA Positivity and VAF Levels Correlate with Macroscopic Residual Disease

Compared with cfDNA in the control group, cfDNA concentrations in patients with hepatoblastoma were measured at high levels with a wide range (45–4992 ng/mL of plasma, median 36 ng/mL; Mann-Whitney test, *p* < 0.0001). CfDNA was dependent on disease stage (localized disease – median 32 ng/mL, metastatic disease – median 57 ng/mL; ordinary one-way ANOVA *p* = 0.0425 and *p* = 0.0005, respectively) (Fig 3a), and the clinical time point it was collected on for samples collected at diagnosis (median 86 ng/mL; ordinary one-way ANOVA *p* = 0.0098, compared with samples collected during neoadjuvant chemotherapy, post-op, or at disease recurrence) (Fig 3b).

**Figure 3.**
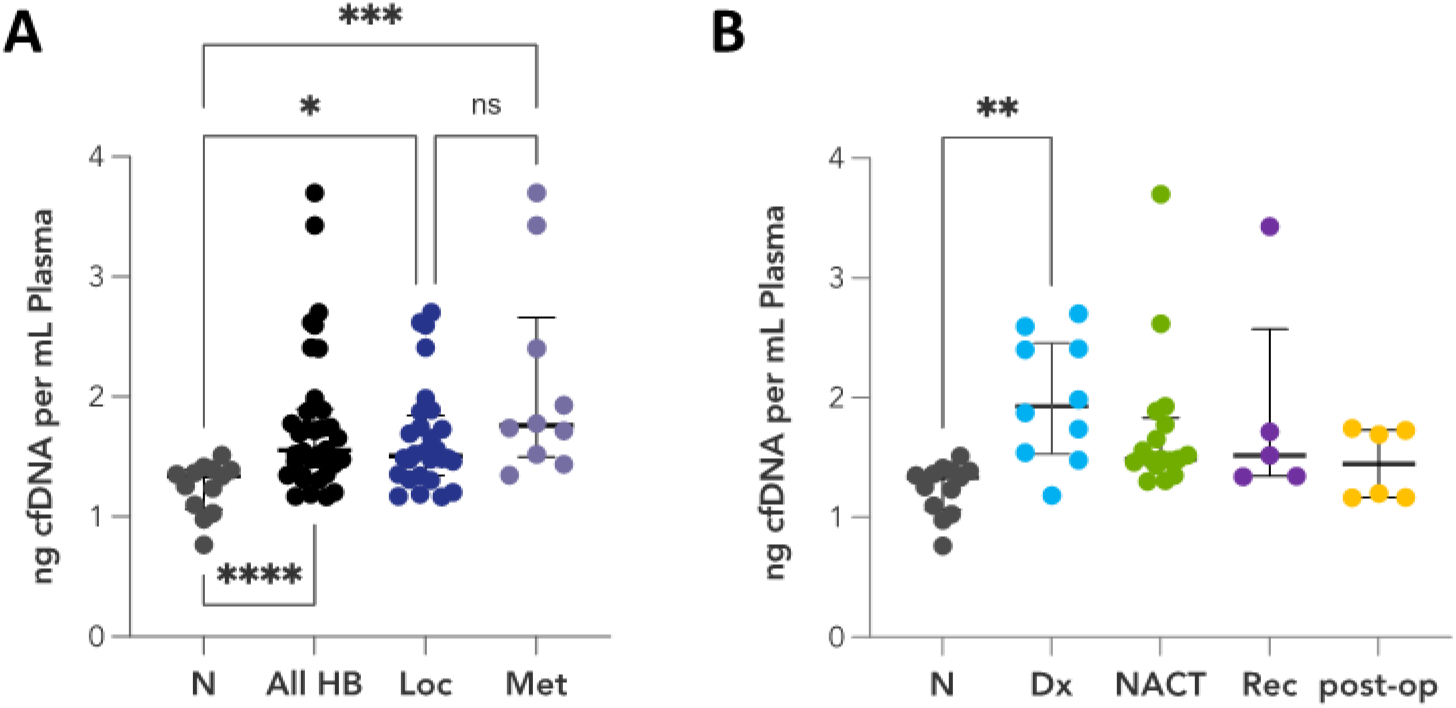
cfDNA levels in hepatoblastoma correlate with disease stage and clinical time point. N – control group, HB – hepatoblastoma, Loc – localized disease, Met – metastatic disease, Dx – diagnosis, NACT – neoadjuvant chemotherapy, Rec – recurrent disease. P-values are presented in asterisks (* =0.0425, ** = 0.0098, *** = 0.0005, **** <0.0001). QUENCH was able to detect variants in 11/17 (65%, 8 SV and 3 SNV) of the Sanger confirmed *CTNNB1*-mutated cases with VAF ranging 0.3-35.4%, including 3 samples (20%) at VAF under 1.5%. NGS *CTNNB1* variant calling was blinded to the independent Sanger sequencing results and all variants were in complete agreement. *CTNNB1* variants were detected in samples taken at initial diagnosis in 9/10 (90%, mean VAF=17.7%, ranging 2.7-35.4%), during/following neoadjuvant chemotherapy in 5/15 (33%, mean VAF=3.5%, ranging 0.3-6.3%), at metastatic recurrence in 1/5 to 1/3 (20%-33%, VAF=1.1%, 3 of the 5 cases with Sanger confirmed *CTNNB1*-mutation), and post-operatively for fully resected localized disease in 0/6 (0%), suggesting a positive correlation between ctDNA positivity and tumor burden (Fig 4a) as well as between VAF levels and tumor burden (Fig 4b).

**Figure 4.**
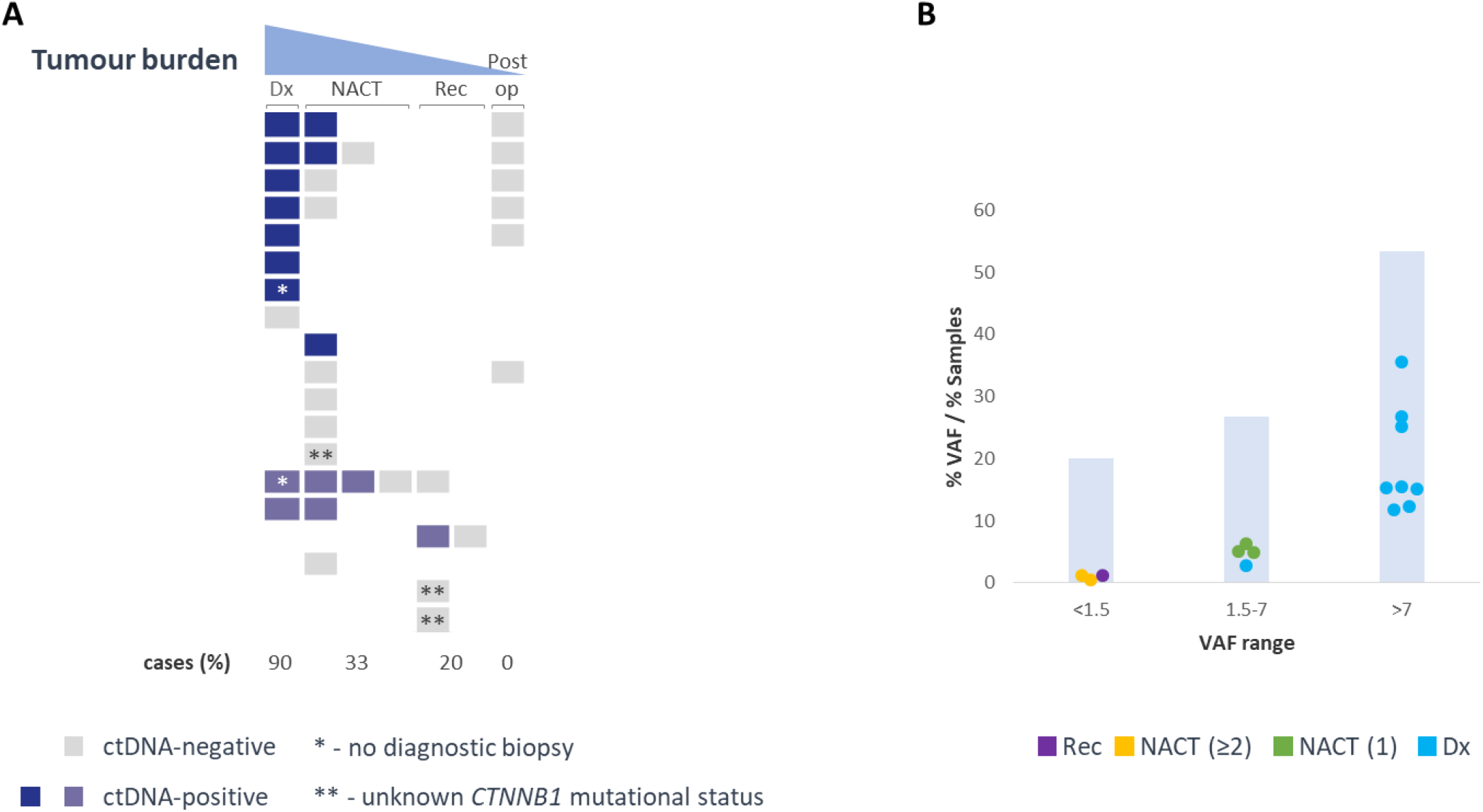
ctDNA positivity correlate with tumor burden (A) and levels correlate with clinical time point (B). **(A)** Each raw represents a different patient. Colored squares represent tested cfDNA samples: grey – ctDNA-negative, indigo – ctDNA-positive in localized cases, purple – ctDNA-positive in metastatic cases. **(B)** Scatter plot of ctDNA-positive samples at different VAF ranges, color coded according to time point of collection. Dx – initial diagnosis – cyan, NACT – during neoadjuvant chemotherapy (the number of previously administered cycles shows in brackets) – green (post 1 cycle) / yellow (post 2 or more cycles), Rec – disease recurrence – purple. CtDNA VAF (for the scatted plot) and % of samples in each VAF range presented in bar chart.

### CtDNA Correlates with AFP

AFP levels were available for 36/38 samples, with abnormal levels detected in 34 samples. Generally, ctDNA levels determined from QUENCH did not correlate with AFP levels, as 17 samples were ctDNA-negative and AFP-elevated. We note that 4 of those samples were taken after a full resection with negative margins of localized tumors. Although the presence of a microscopic residual disease, eliminated by subsequent adjuvant chemotherapy cannot be ruled out, these samples were expected to be negative for hepatoblastoma markers. AFP in these cases fell to the reference range as long as 2 to 9 months postop due to its half-life of 4-9 days ^14–16^.

We thus interrogated the relationship between 15 QUENCH-positive samples and AFP, and found that ctDNA VAF correlated with the order of magnitude of AFP levels (R^2^ = 0.6713, Fig 5a) in all but one sample that was collected from a patient diagnosed with hepatoblastoma at an unusual older age (11-15 years). Although AFP is very sensitive, it showed low specificity compared with both QUENCH and ddPCR (15% Fig 5b, and 17%, Fig 5c, respectively).

**Figure 5.**
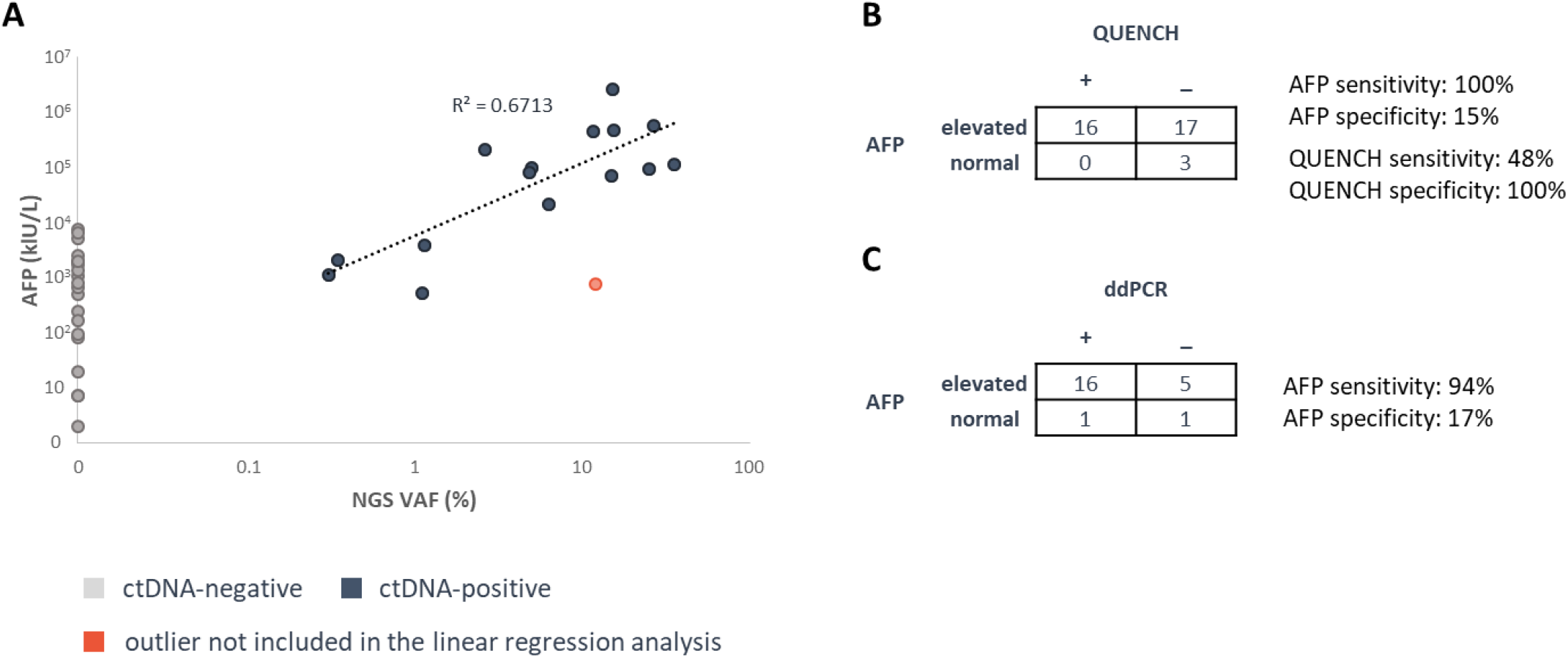
ctDNA correlate with AFP. **(A)** VAF of QUENCH-positive samples and AFP are concordat (R^2^=0.6713). Grey – ctDNA-negative, indigo – ctDNA-positive, salmon – ctDNA-positive outlier not included in the linear regression analysis. Sensitivity, specificity, and concordance of variant detection evaluated by QUENCH and AFP **(B)** and ddPCR and AFP **(C)**.

### AFP and ctDNA Correlate with Tumor Size

Diagnostic MRI or CT imaging studies and cfDNA samples were available for 10 patients, with mean total tumor volume of 780 cm^3^ (range 83–1574 cm^3^). Linear correlation was found between AFP and the volumetric assessment of the tumor size, except for the older patient as in the previous paragraph who remained an outlier here as well (R^2^ = 0.7923, Fig 6a).

**Figure 6.**
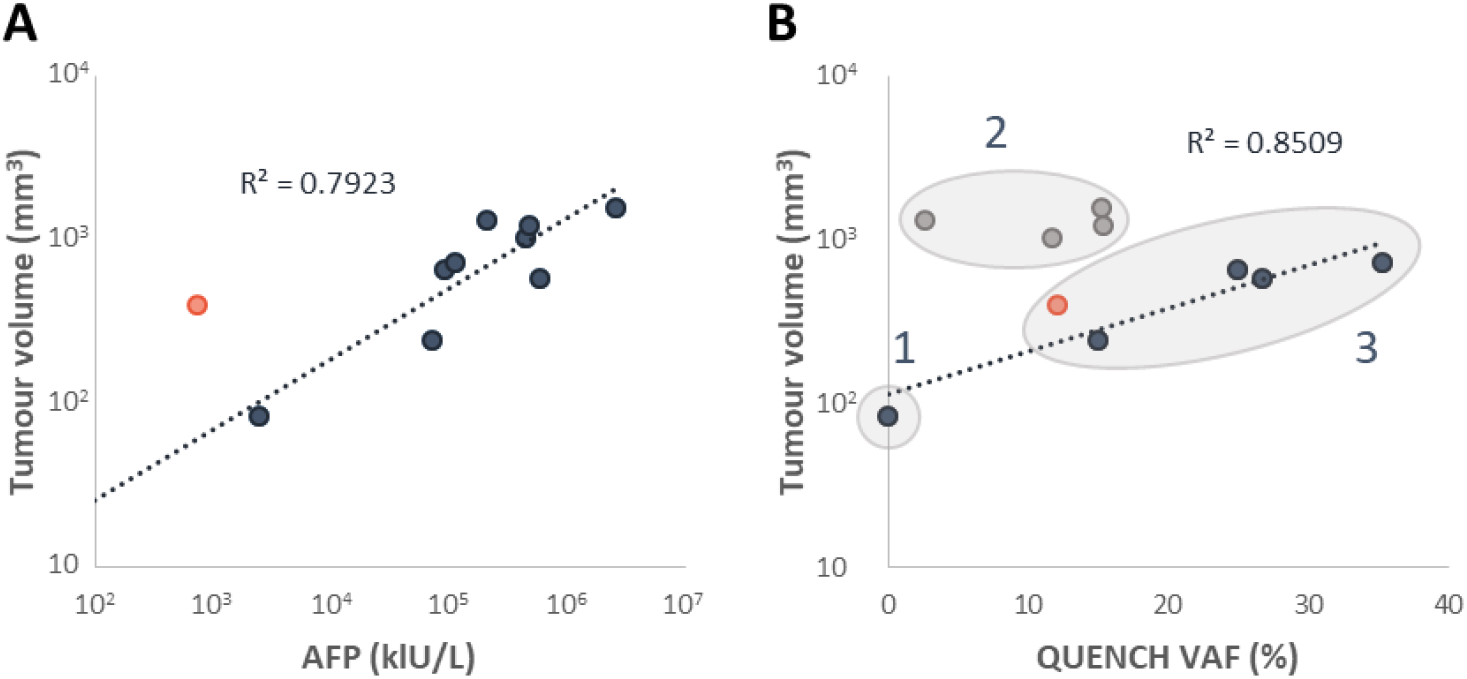
AFP levels correlate with tumor size at diagnosis (A), but only smaller tumors show a correlation between ctDNA VAF and tumor size (B). The relationship between the VAF and tumor size did not hold for Group II, however, we could not identify an association to other clinical, pathological, or radiological parameters (Supplementary Materials Table S1) to explain the different propensity to release ctDNA.

A different pattern was observed for ctDNA. Based on tumor size and ctDNA VAF levels, patients were divided in to 3 groups: Group I with a small tumor (83 cm^3^) and undetectable ctDNA, Group II with moderate ctDNA VAF and markedly large tumors (2.7-15.4%, and 1031-1574 cm^3^, respectively), and Group III with high ctDNA VAF and medium sized tumors (12.1-35.4%, and 245-731 cm^3^, respectively) (Fig 6b). Excluding patients from Group II, a linear correlation was observed between ctDNA VAF and tumor volume (R^2^ = 0.8509, Fig 6b).

### CtDNA VAF Correlate with Dynamic Treatment Response

Longitudinal cfDNA samples (n=23) collected from 7 patients demonstrated similar dynamics of both ctDNA and AFP in 6 patients, reflecting the response to systemic treatment (Fig 7). The only discrepancy was a sample collected at diagnosis which had VAF of only 2.7% (compared with 8 samples collected at diagnosis having VAF 11.7-35.4%). The sequential sample collected at tumor resection had 4.8% VAF, while serum AFP decreased by ∼3 times between these time points. As this sample was collected in EDTA tube, we speculate that this discrepancy is related to pre-analytical conditions of blood collection and/or processing.

**Figure 7.**
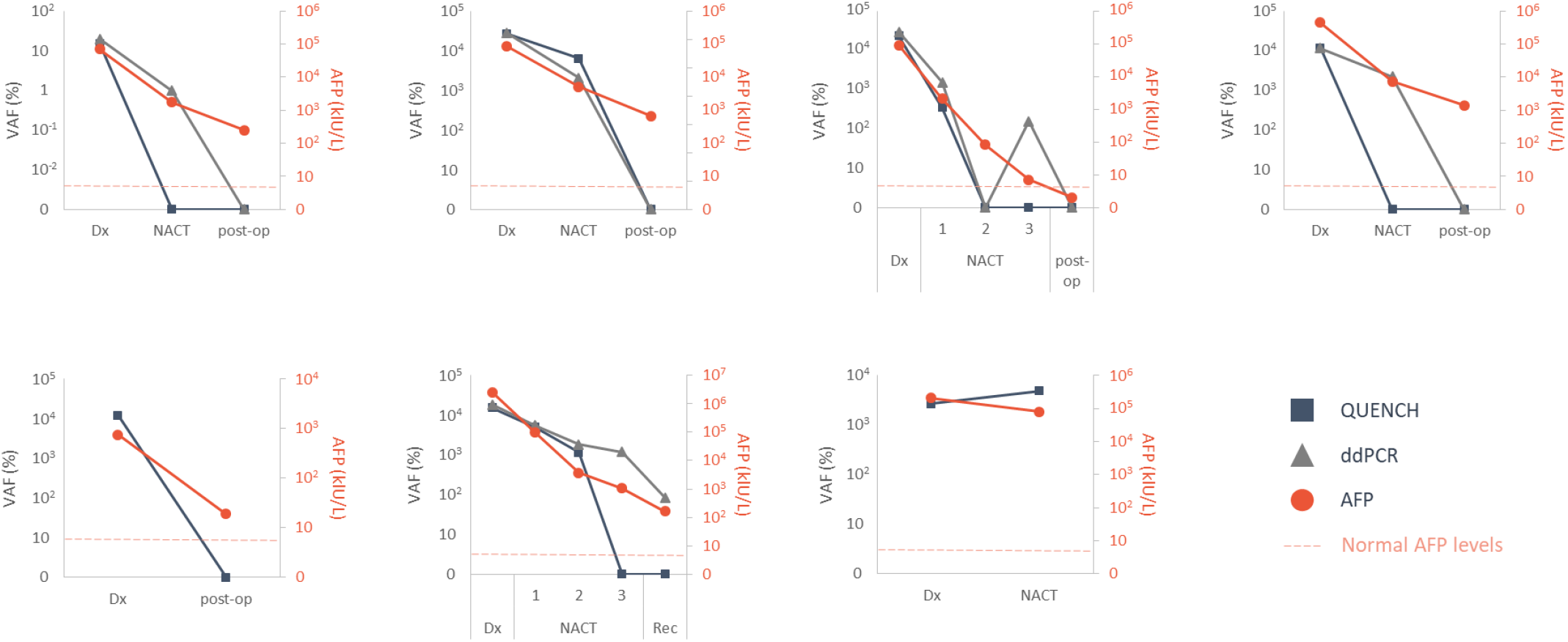
CtDNA correlate with dynamic treatment response. Serial samples with matched ctDNA VAF (evaluated by QUENCH – indigo squares or ddPCR – grey triangles, left Y axis) and AFP levels (salmon, right Y axis), collected at different clinical time points: Dx – initial diagnosis, NACT – during neoadjuvant chemotherapy, post-op – post resection of primary tumor, Rec – disease recurrence. Age-adjusted normal values of AFP presented in dashed line.

Of note, as ctDNA VAF during treatment was low, ddPCR enabled a more refined monitoring to ctDNA changes with 8/9 samples identified through ddPCR at VAF under 2.2% while QUENCH failed to identify 5 of them.

## DISCUSSION

Liquid biopsy is rapidly gaining traction for potentially revolutionizing cancer diagnosis, disease monitoring, and treatment selection through blood-based analysis of shed biomolecules ^8^, circumventing the need for invasive tissue biopsies. In rare tumors like pediatric cancers, the identification of biomarkers with impact on prognosis and treatment remains very challenging. In the context of hepatoblastoma, we have previously demonstrated ctDNA analysis in a small series of 3 cases to be a biomarker for disease monitoring similar to AFP ^12^. However, there are several general as well as hepatoblastoma-specific inherent challenges with a sensitive and quantitative ctDNA assessment:

I. A key requirement for NGS-based ctDNA assays is high sensitivity. Polymerase error during amplification ^17,18^ and sequencing error of NGS platforms ^19,20^ make it difficult to robustly quantitate low-frequency mutations <1% VAF using conventional NGS technologies ^21^. Digital sequencing with UMIs have been developed to suppress the errors to detect variants below 0.1% VAF ^22,23^.
II. However, having deep sequencing coverage alone is not sufficient for detecting variants at very low allele fractions. Importantly, high input DNA amount is needed in the sequencing workflow for VAF calling below 0.1%, which is not feasible for most early- and many advanced-stage cfDNA samples from solid tumors ^24^. Hepatoblastoma arises almost exclusively in young children, with 90% of cases occurring in children less than five years old and primarily before the age of three ^25^. Therefore, the volume of the blood samples that children with hepatoblastoma can provide is limited to usually 1 mL of plasma ^26^, with cfDNA concentrations measured at a median of 36 ng/mL. Typically, about 5,400 diploid human genome equivalents ^13^ are sampled with a maximum sensitivity of 0.02% VAF achieved for 1 variant copy (the minimum requirement for SV variant calling).
III. Approximately half of the variants in hepatoblastoma are large deletions within the *CTNNB1* exon 3 locus (up to about 1000 bp) ^9–11^. CtDNA SVs, including deletions, are not trivial to detect and require several considerations. Firstly, the two principal methods for targeted enrichment of genomic regions for NGS are amplicon-based sequencing that relies on PCR amplification and hybridization-capture that uses complementary probes to capture target sequences. Single-primer extension design is a modified approach to amplicon-based sequencing in which each genomic target is enriched by a target-specific primer and a universal primer. This strategy removes conventional two target-specific primer design restriction and is less dependent on the size of DNA fragments compared with hybridization-capture and so enables capture of ctDNA deletion events in an unbiased manner.

Secondly, the fragmented nature of ctDNA (ranging between 20 and 220 bp, with a maximum peak at 167 bp ^27^) poses a challenge for detection of SVs, as split-read and/or discordant-reads (the regions flanking the break point) are difficult to accurately align with short supporting sequences and require custom analysis.

To overcome these, UMI incorporation as well as single primer extension capture were used in our assay. In addition, an in-house SV calling tool was developed to improve the detectability of *CTNNB1* deletions. Under this framework, LoD of 0.1% VAF and LoB of 0% VAF were determined in clinically relevant DNA input amounts of 20 ng, reflecting 0.5 mL plasma volumes.

A major advantage of this NGS-based method is that it is applicable to nearly all patients with hepatoblastoma, unlike patient-tailored ddPCR assays that require a priori knowledge of the somatic events from tumor tissues and time to design private assays accordingly. Notably, two of the patients in our study had a provisional diagnosis based on imaging and AFP levels, and commenced neoadjuvant chemotherapy to manage their symptoms without histological conformation of hepatoblastoma. *CTNNB1* ctDNA variants were detected in both patients at this time point. However, an additional 2 and 8 months were needed to confirm these variants in the matched tumor samples. Primary tumors were resected 2 and 3 months post neoadjuvant treatment, having adequate tumor material for mutation discovery from only one of them as the other was completely necrotic. Lung metastasis was available 8 months post diagnosis and was used to confirm the second variant.

Serum AFP concentrations are highly elevated in neonates and exceedingly high levels can be observed in premature infants due to their developmental stage ^28^, making AFP an inaccurate measurement in this population. Incidence rate for hepatoblastoma have been increasing in developed countries worldwide in the last decades by about 2.5% per year ^29^, and is attributed in part to the increased survival rates of premature babies, as extremely premature babies with a birth weight of less than 1 kg have been reported to have a greatly increased risk of developing hepatoblastoma ^30^. Taken together, these could impose a difficulty in interpreting AFP values in a bigger proportion of cases in the future.

At present, very little data comparing ctDNA to AFP in hepatoblastoma is available ^12^. One of the major potential advantages of ctDNA testing is that it is tumor-specific and may therefore be a more reliable measure of disease status or the nature of the tumor, while AFP is only tumor-associated. Furthermore, being a driver event, all tumor cells are expected to harbor the *CTNNB1* variant, regardless of tumor heterogeneity. Of note, the only case with undetectable ctDNA at disease diagnosis assessed by QUENCH was also the only patient with very low risk disease. This case however did not have the lowest AFP levels; rather a high-risk tumor presented with the lowest AFP values. The high sensitivity to detect ctDNA at disease diagnosis (9 out of 10 patients, 90%) even for localized cases (7 out of 8 patients, 88%) makes ctDNA liquid biopsy a potentially important tool to refine risk stratification between the very low and low risk groups, although this requires further validation.

We observed an excellent correlation between AFP levels and tumor volume at diagnosis; however, very large tumors did not show an association between tumor volume and VAF levels, implying that additional factors contribute to ctDNA levels in the blood. Interestingly, AFP has also been used to diagnose hepatocellular carcinoma and detect postoperative recurrence for decades, and is facing similar limitations in sensitivity and specificity ^31^. An additional point of note is the correlation of ctDNA VAF with tumor size and AFP levels in hepatocellular carcinoma, as well an association between ctDNA positivity and macrovascular invasion found by Ge Z and colleagues ^32^ in hepatocellular carcinoma. The data for vascular invasion is unfortunately unavailable in our cohort.

We found that ctDNA is a good surrogate marker of tumor burden and have confirmed the capacity of serial ctDNA sampling to monitor dynamic tumor response, similarly to AFP. The ability of QUENCH to accurately quantify VAF was demonstrated by both the high correlation with ddPCR for VAF above 0.3% and by the correlation between VAF levels to the macroscopic residual disease at different clinical time points.

Potential disadvantages of ctDNA over AFP testing include longer turnaround times, increased assay costs, and limited sensitivity at low disease burden. In addition, the implementation of NGS assays into clinical routine use can be limited by the complexity of the associated protocols and data analysis. While QUENCH was able to detect variants in clinical samples down to 0.3%, it had limitations in samples taken at low disease burden. To enable a near real-time method for testing ctDNA, our study employed a tumor-agnostic approach for *CTNNB1* ctDNA detection in which the LoD was determined from the HepG2 variant without patient-specific variant optimizations. Applying to clinical samples with different variants, QUENCH fell short of reliably detecting ctDNA at VAF in the range of 0.1%-2.2% compared with ddPCR. In addition, QUENCH was unable to detect ctDNA in 4/5 samples taken at relapse, and ddPCR in 1/3 of them. Thus, QUENCH may lack the sensitivity required for clinical disease monitoring at low disease burden, such as with MRD.

Two important factors underlying the LoD of all ctDNA profiling methods are the number of cfDNA molecules recovered and the number of mutations in a patient’s tumor that are interrogated ^23^. QUENCH was designed to capture variants in exon 3 of the *CTNNB1* gene as it is a hallmark of sporadic hepatoblastoma, with single-mutation genotyping confining ctDNA detection limits to 0.1% VAF. However, multigene panels assaying many variants instead of a single variant increase the probability of finding detectable variants in plasma samples and provide improved sensitivity relative to a single marker ^23^. For example, Chaudhuri A and colleagues tracked multiple ctDNA variants in patients with stage I-III non–small-cell lung cancer following definitive treatment and reported that a 94% detection rate with multiple markers dropped to 58% using the same platform with only a single marker ^33^. In the last few years it has been recognized that targeted methylated ctDNA analysis demonstrates superior sensitivities and specificities to detect a broad range of cancers, compared with mutation-based ctDNA assay ^34^. Recently, the DNA methylation landscape of hepatoblastoma was profiled to find DNA methylation clusters which tightly correlate with histological subtypes, and clinical behaviors ^11^. In addition, about 600 differentially methylated regions were identified by comparing between hepatoblastoma and their matched non-tumoral liver tissues ^35^. Adding a methylated ctDNA layer to our QUENCH mutation-based ctDNA assay could potentially improve the sensitivity of the detection of low-frequency alleles in ctDNA.

To conclude, changes to AFP values and imaging studies are the current mainstay for treatment monitoring ^36^ and relapse surveillance ^37^ in hepatoblastoma. This study provides promising evidence for the utility of quantitative NGS and ddPCR ctDNA detection as a surrogate marker of tumor burden and treatment response. Since the detection targets are distinct, ctDNA- and AFP-based stratification/monitoring approaches may complement one another, and additional studies are needed to better define the interplay between ctDNA and AFP and the optimal clinical use for ctDNA- and AFP-based MRD methods.

## MATERIALS AND METHODS

### Study Design

The study was conducted under research protocols approved by the Sydney Children’s Hospitals Network Human Research Ethics Committee, and Baylor College of Medicine (reference numbers: HREC/17/SCHN/302, and BCM IRB H-38834, respectively), with informed consent obtained from all participants.

Eligibility criteria included diagnosis of hepatoblastoma, and available plasma samples collected at diagnosis or during treatment and surveillance. Accordingly, 20 patients enrolled, and 38 samples were collected prospectively across the two sites: 25 samples from 9 patients treated at The Children’s Hospital at Westmead (CHW), and 13 samples from 11 patients treated at Texas Children’s Hospital.

An additional thirteen samples were collected at CHW from children in long-term remission from solid cancers (n = 5 neuroblastoma, n = 5 sarcomas, and n = 3 hepatoblastoma) who were otherwise healthy.

AFP serum levels were routinely tested and obtained from the electronic health record. Additional clinical and pathological information can be found in Supplementary Materials: Table S1.

- Samples collected at CHW: 2-3 mL blood samples were collected in 10 mL Streck tubes (Cell-Free DNA BCT®, STRECK, La Vista, NE, USA, catalog No. 218997) and processed for plasma at ambient temperature in a double-centrifugation protocol as previously described ^12^. Plasma aliquots were stored at −80 °C until DNA isolation.
- Samples collected at Texas Children’s Hospital: 2-3 mL blood samples were collected in EDTA tubes and processed for plasma in a double-centrifugation protocol: first centrifugation at 1,200 x g for 10 min followed by plasma supernatant aspiration into new tubes without disturbing the buffy coat layer, then a second centrifugation of the plasma supernatant at 15,000 rpm at 4 °C for 10 min, followed by aspirating the top phase into new tubes without disturbing the pellet, and storing at -80°C until DNA isolation.

### DNA Isolation and Quantification

- Cell free circulating DNA was extracted from 0.5-1 mL of frozen plasma samples using the QIAamp Circulating Nucleic Acid kit (Qiagen, catalog No. 55114, Chadstone, VIC, Australia) according to the manufacturer’s instructions, except for increasing the proteinase digest step to 60 min for plasma samples collected in Streck tubes, as recommended by Streck product literature. DNA was eluted in 40 μL buffer provided with the kit and stored at −80 °C until analysis. DNA quantification was performed using the Qubit dsDNA High Sensitivity Assay Kit for the Qubit 2.0 Fluorometer (Life Technologies, USA).
- Genomic DNA was extracted from cell lines using AllPrep DNA/RNA/Protein kit (Qiagen; catalog No. 80004), and normal peripheral mononuclear cells, and matched whole blood (germline) samples using QIAGEN DNeasy Blood & Tissue kit (Qiagen; catalog No. 69504) according to the manufacturer’s instructions.

### QUENCH

- Library preparation: Cell-free DNA libraries were constructed with a customized QIAseq Targeted DNA Panel Kit (QIAGEN), as described in detail previously ^38^. The input amount preferred for library preparation was 40 ng, but 3.5-40 ng cfDNA samples were included in this cohort (mean 19.4 ng). Briefly, cfDNA was end-repaired, A-tailed, and ligated with unique molecular identifiers (UMIs) barcoded adaptors. The adaptor-ligated libraries were target enriched with PCR using a panel of loci specific primers (8 cycles). The targeted enrichment was performed with a customized QIAseq Targeted DNA Panel primer design to amplify the region covering exons 2-4 of the *CTNNB1* gene. A double-stranded higher tiling density design was used to accommodate for the small cfDNA fragments. The target enriched libraries were further amplified for 23 cycles with PCR and were size selected for an average fragment size of 300 base pairs (bp) (corresponding to insert size of about 110 bp). The library profile was quantified using Qubit dsDNA HS Assay kit (Invitrogen™, Thermo Fisher Scientific, Waltham, MA, USA). The quality and quantity of the prepared library was assessed by the Australian Genome Research Facility (AGRF), Sydney, Australia. The library profile was analyzed with the High Sensitivity D1000 ScreenTape System (Agilent Technologies, Palo Alto, CA) and qPCR with the NEBNext Library Quantification Kit (New England Biolabs Pty Ltd). After quantification, the libraries were normalized and pooled in equimolar quantities. Sequencing was performed with the Illumina MiSeq according to manufacturer’s recommendations using paired-end sequencing (2 × 150 bp) with the MiSeq v.2 reagent kit and a custom primer (Custom Read primer 1) provided with the QIAseq library kit. Median average read coverage was 685 (range 407-3987) and median average UMI coverage of 378 (range 98-1942).
- Data analysis: Raw sequence data in FASTQ format were processed with the QIAseq DNA pipeline available at https://github.com/qiaseq/qiaseq-dna. Briefly, after trimming adapter sequences, reads were mapped to the human reference genome hg19 with BWA MEM, and SNVs/indels were called with smCounter2 ^39^ with default parameters. Read alignments were used for structural variant (SV) analysis using a custom pipeline. Reads were marked as duplicates based on their genomic position and UMI sequence using Picard MarkDuplicates (v2.26.3). Deletions in the *CTNNB1* gene were identified using a custom R software package SVSeq (https://github.com/danshea00/SVSeq). Briefly, candidate deletions were identified as regions between split read segments. In the case of ambiguous sequences mapping to either segment, they were assigned to the first segment. The deletion with the largest number of supporting reads was retained. Deletion VAFs were estimated based on informative reads at each breakpoint (Supplementary Materials Fig S2). For breakpoint I, VAF_i_ = s_i_ / (s_i_ + n_i_) with s_i_ and n_i_ indicating the number of deletion-supporting and non-supporting read pairs, respectively. Reads used for VAF calculation at breakpoint i were required to have read 1, containing the gene-specific primer, map to the non-deleted region flanking breakpoint i. Reads were considered deletion-supporting or non-supporting depending on whether read 2 mapped to the non-deleted region flanking the second breakpoint, or the deleted region, respectively. Read overlap was required to be at least 19 bp, the seed length used for BWA-MEM alignment. A final VAF estimate was obtained as the weighted average of the breakpoint estimates VAF = w_1_ VAF_1_ + w_2_ VAF_2_ with w_i_ = t_i_ / (t_1_ + t_2_) and t_i_ = n_i_ + s_i_.

**Figure S2.**
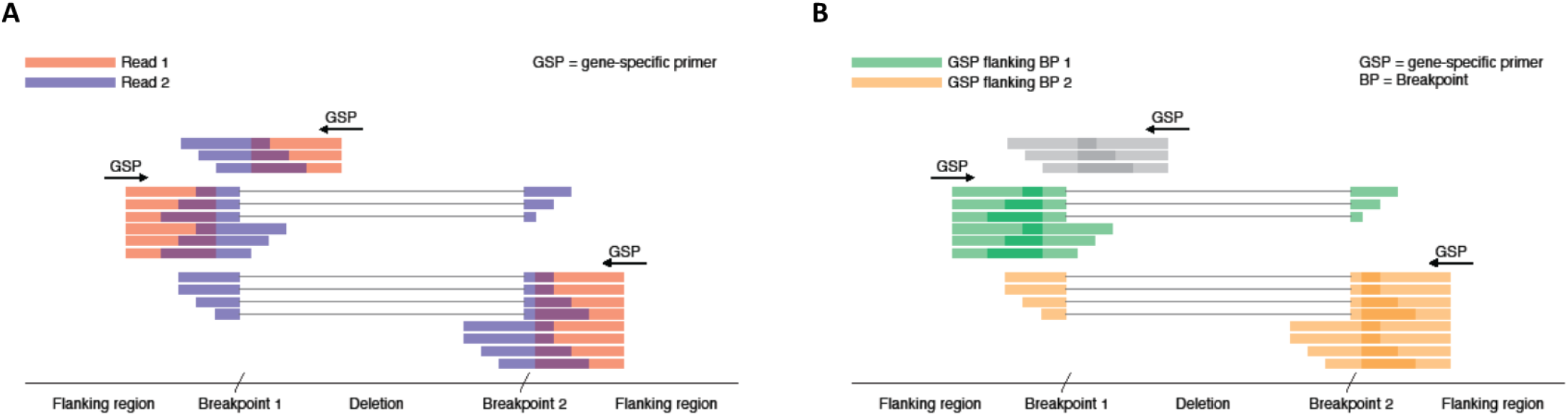
Schematic illustration of VAF estimation. **(A)** Paired reads mapped to a genomic region with a deletion. Red and blue indicate the first (read 1) and second read (read 2) in a pair, respectively. Read 1 includes the gene-specific primer (GSP) and maps to defined genomic positions targeted by the GSP. Shown are reads for three distinct GSPs. **(B)** Reads as in (A) shown in different colors for different GSPs. Colors indicate whether the GSP was (1) flanking the deletion upstream of breakpoint 1 (green), (2) flanking the deletion downstream of breakpoint 2 (orange), or (3) targeting the deleted region (grey). Reads (1) and (2) are used for estimating VAFs at breakpoint 1 and 2, respectively.

### QUENCH limit of detection (LoD)

DNA from the hepatoblastoma cell line HepG2, harboring the 548 bp deletion CTNNB1:c.73_420del ^40^, was mixed with a wild type *CTNNB1* genome to produce artificial VAF levels of 0%, 0.03%, 0.1%, 0.2%, 0.3%, 1%, 1.2%, 5%, 25%, and 50%. We used clinically relevant DNA input amount of 20 ng (see Results) and sequenced to a median average read depth of 627x (407-837x) and median average UMI depth of 307x (221-430x). Deeper sequencing of libraries with shallow read depth did not result in new detections, suggesting sensitivity was limited by library complexity.

### QUENCH limit of blank (LoB)

LoB is the highest mutated *CTNNB1* VAF calling expected to be found in samples containing mutation-negative *CTNNB1* ^41^. To provide QUENCH false-positive call rate, 20 ng input DNA from 4 samples with a wild type *CTNNB1* were used: cfDNA sample from a patient with hepatoblastoma (Supplementary Materials: Table S1), and the three cell lines A673, ES8, and Kelly.

### ddPCR

Mutant (MT) and wild type (WT) *CTNNB1* sequences were used for designing ddPCR (Bio-Rad Laboratories) assays following the dMIQE guidelines ^42^ (Supplementary Materials: Table S2). ddPCR reactions were assembled using standard protocol as previously described ^12^: ddPCR reaction consisted of 10 μl ddPCR™ Supermix for Probes (No dUTP) (Bio-Rad Laboratories), 900 nM/reaction of primers mix for both MT and WT *CTNNB1*, 250 nM/reaction of probes mix for both MT and WT *CTNNB1*, 4 units of restriction enzyme (HaeIII or MseI, New England BioLabs, as per Supplementary Materials: Table S2), for a final volume of 20 μl. CfDNA was added at the same amount as was used for the QUENCH library preparation. Non-template controls (NTCs) contained purified water instead of cfDNA. Tm was optimized for each of the assays using gradient PCR on matched tumor DNA. Assays specificity was determined on matched whole blood (germline) genomic DNA as well as genomic DNA from 2 to 5 different pediatric cell lines (data not shown).

The ddPCR reaction mixture was used for droplet generation, and ddPCR was performed using the QX200 ddPCR system according to manufacturer’s instructions (Bio-Rad Laboratories). QuantaSoft™ Analysis Pro v1.0 software (Bio-Rad Laboratories) was used for data analysis. Each sample was tested in 2 to 3 replicates preformed in at least 2 different experiments.

HepG2 CTNNB1.p.W25_I140del mutation status was confirmed by Sanger sequencing. The relative copy number of both MT and WT alleles was determined by comparing to a normal genome from a healthy individual (data not shown).

### Statistical Analysis

Linear regression was performed using Excel Microsoft Office Professional Plus 2013. Mann– Whitney nonparametric t test, and ordinary one-way ANOVA were performed using GraphPad Prism version 9.3.1 for macOS (GraphPad Software, San Diego, CA, USA, www.graphpad.com). Sensitivity was calculated as the fraction of positive samples that were classified as positive by the alternative method. Specificity was calculated as the fraction of negative samples that were classified as negative by the alternative method.

## Data Availability

All data produced in the present work are contained in the manuscript.

## Data and materials availability

All data are available in the main text or the supplementary materials.

## Acknowledgments

We thank the patients and parents for their contribution to this study. The authors thank the Sydney Children’s Tumour Bank Network (Children’s Hospital at Westmead Tumour Bank: A/Prof Daniel Catchpoole, Aysen Yuksel, Dr Li Zhou, and Natalie Gabrael) and Dr Nicole Graf from the Pathology Department at the Children’s Hospital at Westmead for providing tumor samples. The HepG2 cell line kindly provided by A/Prof. Liang Qiaoand (The Westmead Institute for Medical Research, NSW, Australia). Peripheral mononuclear cells from a healthy individual kindly provided by the Australian Red Cross Blood Service (NSW, Australia). Neuroblastoma cell line Kelly purchased from ECACC (European Collection of Authenticated Cell Cultures), sarcoma cell lines – A673, and ES8 kindly provided by Dr Belinda Kramer (Advanced Cellular Therapeutics, Children’s Cancer Research Unit, Kid’s Research, The Children’s Hospital at Westmead, NSW, Australia). ddPCR was performed at the Westmead Scientific Platforms, which are supported by the Westmead Research Hub, the Cancer Institute New South Wales, the National Health and Medical Research Council and the Ian Potter Foundation. AGRF is supported by the Australian Government National Collaborative Research Infrastructure Initiative through Bioplatforms Australia.

## Funding

This research was funded by the Tour de Cure grant RSP-191-2020 (JK, SKE, SAV), U.S. Department of Defense Career Development Award grant CA201061 (SEW), and Cancer Prevention and Research Institute of Texas (CPRIT) Multi-Investigator Research Award grant RP180674 (SAV). SKE salary was supported by the Royal Australasian College of Surgeons (RACS) grant 170287 (JK). JT, DTPS, AEM, SK, and LDG acknowledge funding from the Kinghorn Foundation.

## Disclosure and Competing Interests Statement

Authors declare that they have no competing interests.

## Author contributions

Conceptualization: SKE, JK

Sequencing data analysis and software development: JT, LDG, AEM, DPTS

Sequencing and ddPCR design, data generation, statistical analysis: SKE

Samples acquisition and clinical data curation: LEC, AM, GM, SEW, SAV, JK, SKE

Visualization: SKE

Writing – original draft: SKE

Writing – review & editing: LDG, LEC, SEW, JT, JK

Supervision: JK, SAV, GM

## Figure legends

**Figure S1 - Schematic view of the *CTNNB1* exon-intron structure targeted by QUENCH and the mutations in the studied cohort and HepG2 cell line.** Pathogenic SNV missense mutations (in circle - the number of patients with the same mutation) and SV deletion mutations identified in the study cohort and in the HepG2 cell line are shown.

**Figure S2 - Schematic illustration of VAF estimation. (A)** Paired reads mapped to a genomic region with a deletion. Red and blue indicate the first (read 1) and second read (read 2) in a pair, respectively. Read 1 includes the gene-specific primer (GSP) and maps to defined genomic positions targeted by the GSP. Shown are reads for three distinct GSPs. **(B)** Reads as in (A) shown in different colors for different GSPs. Colors indicate whether the GSP was (1) flanking the deletion upstream of breakpoint 1 (green), (2) flanking the deletion downstream of breakpoint 2 (orange), or (3) targeting the deleted region (grey). Reads (1) and (2) are used for estimating VAFs at breakpoint 1 and 2, respectively.

## List of Supplementary Materials

Figure S1 - Schematic view of the CTNNB1 exon-intron structure targeted by QUENCH and the mutations in the studied cohort.

Figure S2 - Schematic illustration of VAF estimation.

**Table S1.**
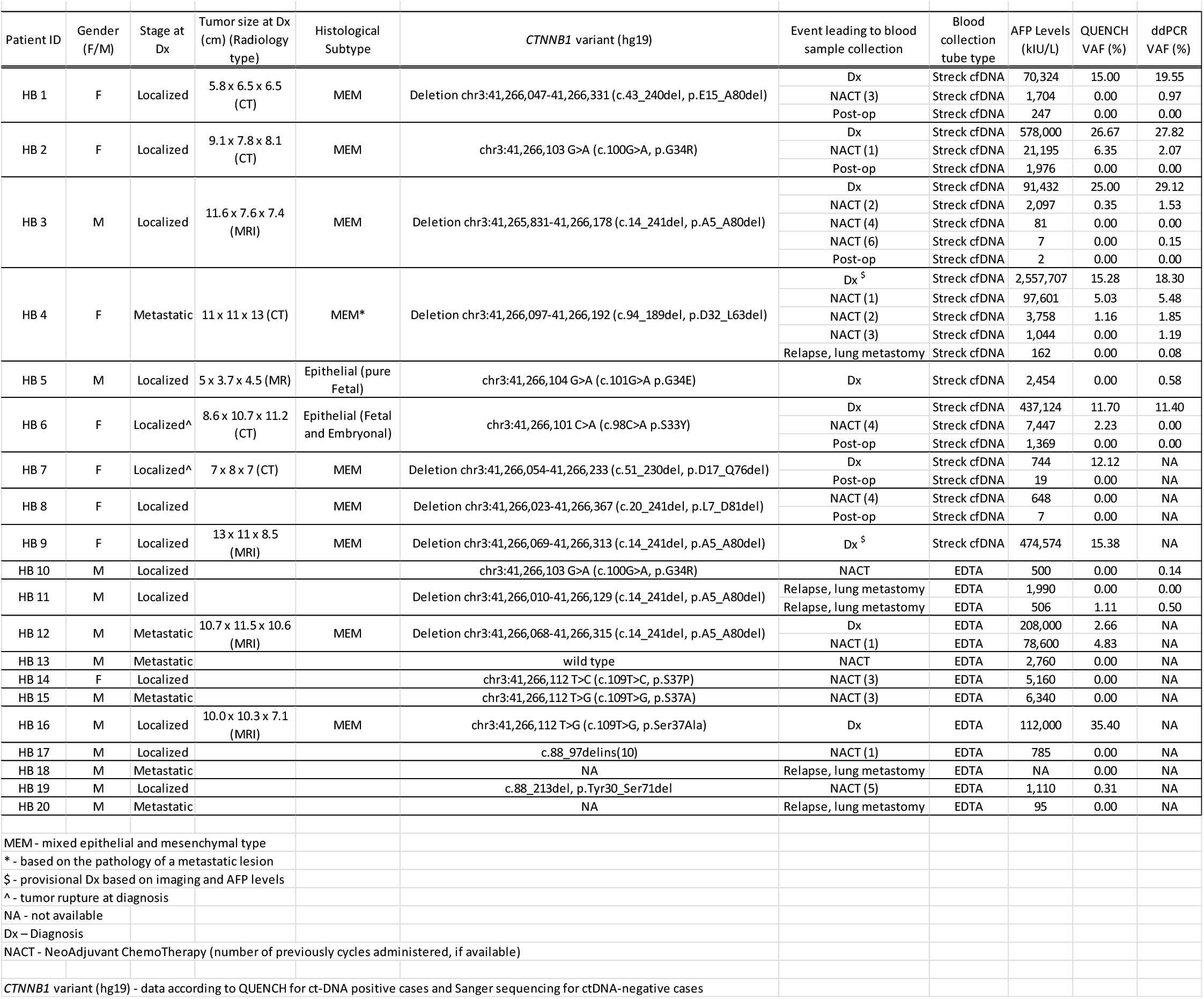
Clinicopathological data and mutational status of the study cohort.

**Table S2.**
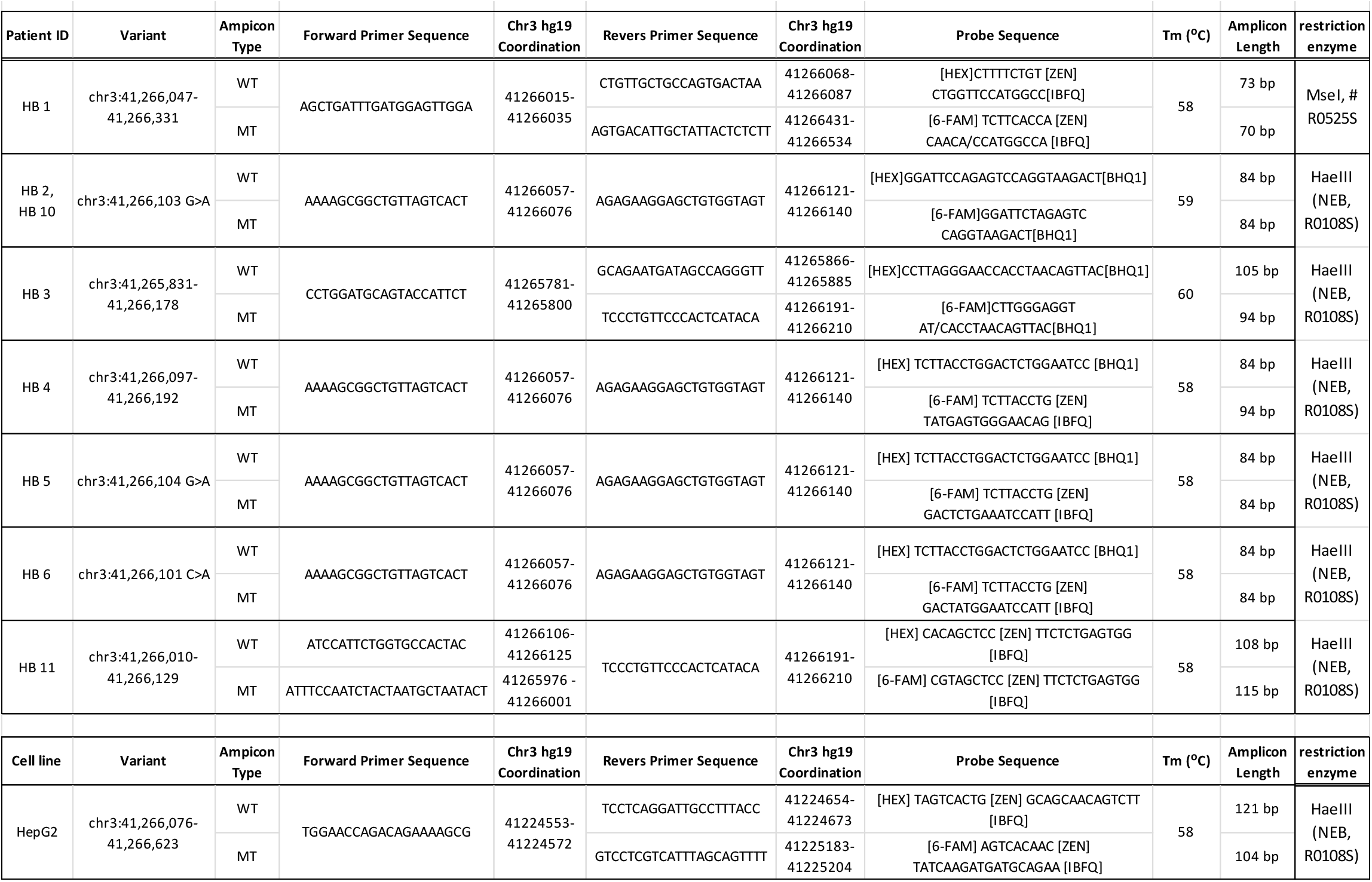
ddPCR primers/probes sets information.

## Notes

### Competing Interest Statement

The authors have declared no competing interest.

